# Academic Achievements in Adolescents with Congenital Heart Disease: A Total Population-Based Cohort Study

**DOI:** 10.64898/2026.01.06.26343567

**Authors:** Sofia Ekmark-Sergel, Michael Lundberg, Cecilia Magnusson, Ulrika Ådén, Gunnar Bergman, Veronica Siljehav

**Affiliations:** Department of Women’s & Children’s Health, Karolinska Institutet, S-171 76 Stockholm, Sweden; Department of Global Public Health, Karolinska Institutet, S-171 77 Stockholm, Sweden; Centre for Epidemiology and Community Medicine, Region Stockholm, S-104 31 Stockholm, Sweden; Department of Biomedical and Clinical Sciences, Linköpings University, S-581 83 Linköping, Sweden; Department of Pediatric Cardiology, Stockholm-Uppsala, Karolinska University Hospital, S-171 64 Stockholm, Sweden

**Author notes:** Correspondence to: Veronica Siljehav, MD, PhD, K6 Women’s and Children’s Health, Karolinska Institutet, 171 77 Stockholm. G. Bergman and V. Siljehav contributed equally.

## Abstract

**Background:** Improved survival in children with congenital heart disease (CHD) has unveiled associated long-term neurocognitive impairments. However, the long-term effects on academic performance and the influence of sex, family, socioeconomic factors, and surgical era remain understudied, limited by small and incomplete cohorts.

**Methods:** This total-population cohort study, with nested sibling analysis, included 1 800 477 singletons born in Sweden between January 1, 1987, and December 31, 2005. Academic achievements at age 16 were assessed by upper secondary school eligibility, total grade sum, and subject-specific grades. Poisson and logistic regression were used to estimate adjusted prevalences, risk ratios (RR), and risk differences (RD). Adjustments were made for year of birth and socioeconomic status. Stratification by noncardiac congenital anomalies, sex, and surgical era was performed.

**Results:** Among 1 800 477 Swedish-born singletons with complete socioeconomic data and residing in Sweden at 16 years of age, 16 075 (0.9%) individuals were identified with CHD and classified hierarchically as univentricular heart (UVH, n=349), severe (n=1 939), moderate (n=1 764), or mild CHD (n=12 023). After excluding major noncardiac congenital anomalies, the adjusted RD for failing to meet upper secondary school eligibility compared to children without CHD was 11 (95% confidence interval 5 to 17), 7 (4 to 9), 6 (3 to 8), and 2 (2 to 3) additional children per 100 for UVH, severe, moderate, and mild CHD, respectively. Children with both CHD and noncardiac anomalies had poorer academic outcomes than those with CHD alone. Additional risk factors were small for gestational age RR 2.34 (95% confidence interval 2.04-2.69) and prematurity RR 1.81 (95% confidence interval 1.64-1.99). Findings were consistent across core subjects, total grade sum, in sibling analyses, and over time.

**Conclusion:** Adolescents with CHD inherently face poorer academic performance, worsening with disease severity and persisting over time. This highlights ongoing educational disparities, especially among children with complex and univentricular heart disease.

**What Is New?:**

- This nationwide cohort study demonstrates that academic achievement declines with increasing congenital heart disease (CHD) complexity, with disparities persisting after adjustment for socioeconomic and familial factors.
- Small for gestational age is identified as an independent risk factor for reduced academic outcomes in children with CHD, a previously unrecognized association.
- Despite the advancements in surgical survival rates over recent decades, academic outcomes did not demonstrate similar improvements.

**What Are the Clinical Implications?:**

- Multidisciplinary follow-up should extend beyond cardiac care in children with CHD, to include cognitive and educational support.
- Small for gestational age status represents an additional risk factor and should be incorporated into risk stratification.
- Genetic abnormalities associated with CHD significantly affect academic outcomes; routine genetic testing should be considered to guide individualized care.

## Introduction

Congenital heart disease (CHD) is the leading major congenital malformation, accounting for nearly one-third of all cases^1^. Advances in pediatric cardiac surgery have been successful, and today, roughly 97% of all children with congenital heart disease are expected to survive into adulthood^2^. This marks a dramatic improvement, as survival rates in the 1980s were only around 85%. The success is greatly attributed to novel surgical techniques and improved perioperative care. In Sweden, the improved survival coincided with the centralization of pediatric heart surgery in the mid 1990s^3^. With increasing survival rates and the larger portion of individuals with CHD now being adults, it has become apparent that they face an increased risk of neurodevelopmental impairments already in childhood^4^. This has been highlighted as one of the most significant morbidities affecting their quality of life both in the short and long term^5-7^. The cognitive challenges in children with CHD are believed to result from a range of factors, including the complexity of the heart disease and concomitant surgery, genetic predisposition, potential complications following surgeries or medications, and socioeconomic factors^4^. Brain dysmaturation is present already in fetal life, due to altered fetal circulation and associated fetal cerebral oxygen consumption and has been linked to adverse neurodevelopment^8,9^. Their brain development can be further affected postnatally by altered brain hemodynamics and white matter damage, resulting from both the heart defect itself and the timing, duration, and number of surgeries required for correction^10-12^. Consequently, the cumulative disruptive effects on brain maturation may impair cognitive development, which, in turn, can impact school performance.

Understanding the impact of CHDs on school achievement is essential, as educational outcomes are well-established predictors of long-term quality of life, mortality, and overall health^13,14^. Besides reflecting knowledge and cognitive ability, grades also reflect broader adaptive functioning^15^ and are influenced by socioeconomic factors^16^. Socioeconomic status also influences neurodevelopmental outcomes in children with CHD^17^ and is considered a risk stratifier^4^; accordingly, it has been emphasized that studies on school performance in children with CHD must account for it^18^. Previous studies on early childhood academic performance with CHD are limited, and cognitive deficits may not yet be evident^19,20^. Many have been unable to adjust for socioeconomic status and familial confounding or have suffered substantial attrition due to non-response, loss to follow-up, or missing data from private schools^18,21-25^. Univentricular heart (UVH), which represents the most neurodevelopmentally vulnerable CHD subgroup^26^, has yet to be delineated in national cohorts^21,22^. Additionally, it has not been shown whether increased survival rates also improve long-term cognitive outcomes^2^.

This population-based cohort study aimed to investigate academic outcomes in adolescents with varying levels of CHD complexity. Among 1.8 million Swedish-born singletons residing in the country at age 16, we identified 16 075 (0.9%) adolescents with CHD. Risk stratification followed the American Heart Association’s (AHA) recommendations, incorporating surgical procedures and their timing.

We assessed school grades and eligibility for higher education, adjusting for socioeconomic factors, accounting for familial confounding, and stratifying by surgical era to clarify the impact of CHD on educational attainment.

## Methods

### Data sources

Using the unique personal identity that is assigned to all Swedish residents^27^, we linked data between multiple registers. Those provided by the Swedish National Board of Health and Welfare were: the Swedish Medical Birth Register (MPR), which has almost complete coverage of all births since 1973 and improved quality since 1982^28^ and the National Patient Registry (NPR), which includes nationwide information on diagnoses for inpatient care since 1987 and hospital outpatient care since 2001^29^. Those provided by the government agency Statistics Sweden were: the Total Population Register^30^ with information on country of birth, death and migration, The National School Registers^31^, where data on graduation grades are routinely collected in the Registry of Compulsory School Leaving grades and the Longitudinal Integrated Database for Health Insurance and Labor Market Studies^32^ providing information on parental education and income. The study was conducted in accordance with the Helsinki Declaration and approved by the Regional Ethical Review Board in Stockholm, Sweden, DNR: 2020-05516, 2021-05958-02, 2022-05648-02 and 2024-00060-01. According to current Swedish regulations, no informed consent is required for research using national registry data.

### Study population

This population-based cohort study included 1 938 397 births recorded in the MBR from the 1^st^ of January 1987 to the 31^st^ of December 2005. A flow chart for inclusion and exclusion is shown in **Figure S1.**

### Congenital heart disease

Diagnoses of CHD and compatible surgery or intervention procedure codes were retrieved from the NPR using the International Classification of Diseases 9 and 10 (ICD-9, ICD-10) and the Classification of surgical procedures (K06 and KVÅ/KKÅ)^33^. Following the New England Regional Infant Cardiac Program^34^, a coding hierarchy was used to classify CHD, ensuring each lesion complexity was assigned to one, and only one of 69 designated hierarchical numbers. Although not exhaustive, the list of included diagnoses represents the most common types of CHD. These diagnoses were further grouped based on complexity, considering the risk of adverse neurodevelopmental outcomes as outlined in the 2024 Scientific Statement from the AHA^4^.

Diagnoses of CHD were obtained before the expected attainment of final grades from compulsory school at age 16. For the UVH group, a diagnosis aligned with single ventricle and a surgical procedure code consistent with at least step 2 of 3 in Fontan palliation was required^35^. The surgical procedure code alone for UVH was also accepted, but the primary diagnoses of the cases were reviewed separately to ensure accuracy. For the severe CHD group, a surgical procedure code matching the CHD or the procedure code of having been treated with cardiopulmonary bypass was required within the first year of life. For children classified as moderate CHD, a surgical procedure code aligned with the CHD was required before the expected attainment of final grades. For the mild unoperated group (mild CHD), all surgical procedure codes for heart surgery (Chapter 3 in K06 and Chapter F in KVÅ/KKÅ^33^) were used to omit surgical cases, ensuring only non-surgical cases were included. A list of all included CHD diagnoses, organized hierarchically and grouped by complexity based on compatible surgical or interventional procedure codes, is provided in **Table S1**.

Children with patent ductus arteriosus associated with prematurity or acquired mitral insufficiency due to rheumatic disease were identified using ICD-9 or ICD-10 and removed from the CHD group.

Children in the severe CHD group expected to have surgery within the first year of life but did not were omitted in the CHD cohort, including: common arterial trunk, aortopulmonary septal defect, double outlet right ventricle, atrioventricular septal defect, total anomalous pulmonary venous drainage, pulmonary atresia with intact ventricular septum, interrupted aortic arch, and tetralogy of Fallot. They were removed to prevent misclassification and align with current pediatric surgical practices.

Non-cardiac congenital anomalies (CA) were identified using ICD-10 (Q00-Q99) and ICD-9 (740-759). Major non-cardiac congenital anomalies (MCA) excluded CHD ICD-10 (Q20-Q26) and ICD-9 (745-747) and minor non-cardiac congenital anomalies as defined by the Swedish National Board of Health and Welfare^33,36^ and presented in **Table S2**.

### Main outcome measures

Qualification for Upper Secondary School (USS) in children without MCAs was our primary outcome, which consists of a composite of grades in English, Swedish, and Mathematics, along with an additional five subjects from the 9^th^ year of compulsory school in the Swedish school system.

The outcome variable comprised three categories: 1) qualification yes or 2) no, or 3) if the student had not completed the last year of compulsory school, leading to non-reported grades (neither pass nor no pass in any subject). The latter is applicable for school dropouts or students with intellectual disabilities requiring special schooling. Not having grades reported in the system has previously been used for imputation as having grades within the lowest 1^st^ percentile^37^.

In a sub analysis, we evaluated the risk of receiving only a passing grade rather than a higher grade in the core subjects and the total grade point, which comprises the sum of all grades^31^, to gain a more nuanced assessment of the child’s scholastic performance. All Swedish schools, whether public or private, are required to report grades to the Swedish National Agency for Education. We imputed missing grades as no grades^37^. An overview of the Swedish school system is provided in **Figure S2.**

### Covariates

We evaluated 2 main categories of potential factors associated with academic performance:

1. Child demographic characteristics such as gender^38^ and year of birth^39^.
2. Sociodemographic characteristics^17,40^ (highest education of parents by the time of birth, country of birth for parents, disposable income/family member at time of birth, maternal and paternal age by birth).

Additional clinical risk factors attributed to CHD and possibly affecting neurodevelopmental outcome include non-cardiac CAs^41^, gestational age^21^, and relative birthweight^17^ (calculated as the centile of birth weight within the given gestational age^42^). However, to avoid over-adjusting for gestational age (GA) and small for gestational age (SGA), which are causal pathways for CHD, they were analyzed separately in relation to their impact on the outcome in children with CHD.

A secondary analysis was performed to elaborate the risk factor of being born before or after the centralisation of pediatric heart surgery (1992-1996)^3^, stratified on period by birthyear. This was merely exploratory since survival significantly increased after centralisation^3^.

### Exclusion

We excluded multiple births (*n*=53 197), children who emigrated (*n*=37 854), or died before graduation at the age of 16 (*n*=10 845). There were no missing data on children’s or parents’ personal identification numbers, and no missing information on sex. Missing socioeconomic data (*n*=36 025) led to excluding cases that could not be adjusted. Additionally, all children with pulmonary atresia with ventricular septal defect and major aortopulmonary collateral arteries (*n*=19) were excluded, as they represent a distinct group with high morbidity but with too few cases.

### Statistical Analysis

All analyses were conducted by using SAS (version 9.4). One author (M. L.) had full access to all study data and takes responsibility for its integrity and analysis. This article conforms to the STROBE (Strengthening the Reporting of Observational Studies in Epidemiology) guidelines for observational studies. We used multivariable Poisson regression models with robust variance to estimate adjusted risk ratio (RR) (with 95% confidence interval (CI)) for each academic outcome by the status of CHD. To estimate Risk Differences (RD), an identity link was added to the model. RD are presented in percentages (%). Total grade sum was evaluated by multiple regression models within the framework of general linear models. In addition to the status of CHD, the models included adjustments for covariates such as year of birth, parents’ highest education at the time of birth, country of birth for parents, family income (disposable income/family member), and maternal and paternal age at birth. Major and minor congenital anomalies were either excluded or retained in the analysis. Children were stratified by sex, GA, and relative birth weight.

### Sensitivity Analysis

To evaluate confounding bias from school dropouts and children in special schools unable to receive final grades, we investigated the effect of adjusting the inclusion of children without information on grades by imputing them as not having passed grades in the subjects with missing information, grade sum=0^37^. The group of children with CHD had proportionally fewer grades compared to the total population.

Finally, shared family factors, as well as unmeasured factors (e.g., parental genetic or unmeasured social factors), were considered using sibling analysis, since siblings share 50% of genetics^43^. In a subset of families with more than one child, we matched each child not eligible for higher education (cases) with one or more full siblings eligible for higher education (controls). To compare the risk of non-eligibility in children with congenital heart disease with that of their eligible siblings, we estimated Odds Ratios (ORs) in conditional logistic regression models matched on family identification number, adjusted for non-shared confounding characteristics.

## Results

Among 1 938 397 liveborn children in Sweden between 1987 and 2005, 1 800 477 were retained in the current study, identifying 16 075 individuals with a congenital heart disease, where 349 (0.02%) were UVH cases, 1 939 (0.1%) were severe CHD, 1 764 (0.1%) were moderate CHD and 12 023 (0.7%) were mild, non-surgical cases. **Figure S1** shows flowchart of inclusion/exclusion. The sum of missing information accounted for 2.0% and was due to incomplete information on socioeconomic factors.

Among children with CHD, 32.2% had a concomitant MCA and 39.8% had any CA, compared with the total population excluding CHD, where 5.5% had an MCA and 10.2% had any CA, as described together with baseline characteristics in **Table 1**. In total, 254 670 children were identified in the subset of families with at least one sibling ineligible for USS and included in the sensitivity analysis.

**Table 1.**
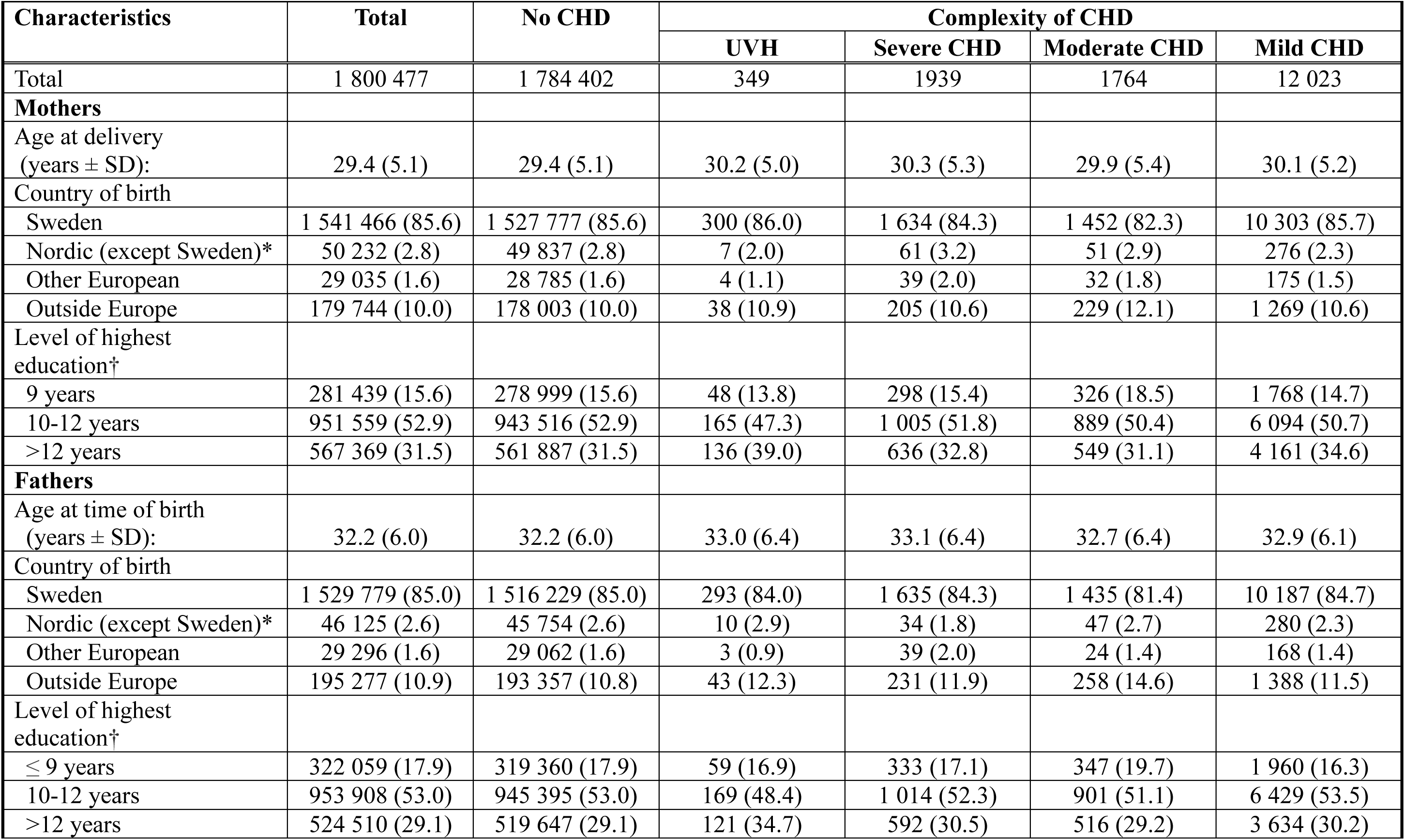
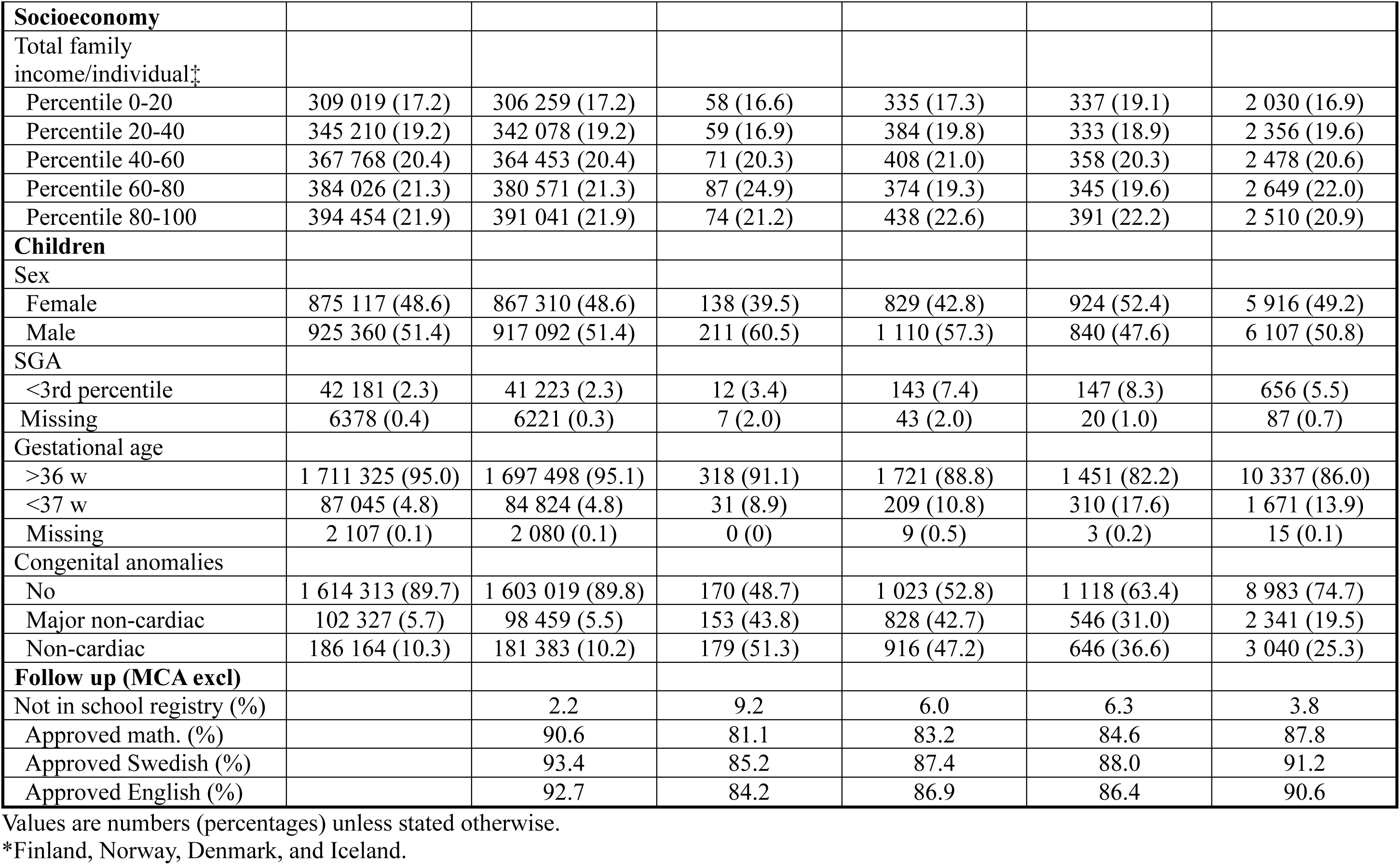
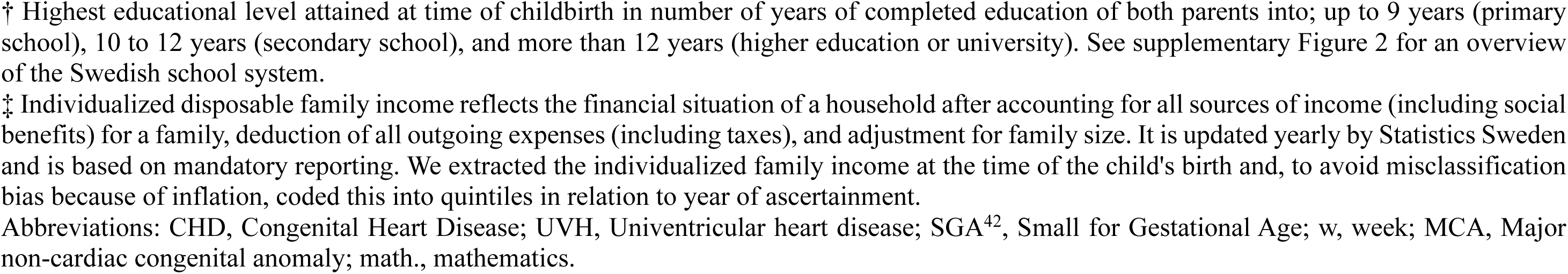
Characteristics of parents and their singletons born in Sweden, 1987-2005, with or without CHD, and residing in Sweden at age 16.

| Characteristics | Total | No CHD | Complexity of CHD |  |  |  |
| --- | --- | --- | --- | --- | --- | --- |
|  |  |  | UVH | Severe CHD | Moderate CHD | Mild CHD |
| Total | 1 800 477 | 1 784 402 | 349 | 1939 | 1764 | 12 023 |
| **Mothers** |  |  |  |  |  |  |
| Age at delivery (years ± SD): | 29.4 (5.1) | 29.4 (5.1) | 30.2 (5.0) | 30.3 (5.3) | 29.9 (5.4) | 30.1 (5.2) |
| Country of birth |  |  |  |  |  |  |
| Sweden | 1 541 466 (85.6) | 1 527 777 (85.6) | 300 (86.0) | 1 634 (84.3) | 1 452 (82.3) | 10 303 (85.7) |
| Nordic (except Sweden)* | 50 232 (2.8) | 49 837 (2.8) | 7 (2.0) | 61 (3.2) | 51 (2.9) | 276 (2.3) |
| Other European | 29 035 (1.6) | 28 785 (1.6) | 4 (1.1) | 39 (2.0) | 32 (1.8) | 175 (1.5) |
| Outside Europe | 179 744 (10.0) | 178 003 (10.0) | 38 (10.9) | 205 (10.6) | 229 (12.1) | 1 269 (10.6) |
| Level of highest education<sup>†</sup> |  |  |  |  |  |  |
| 9 years | 281 439 (15.6) | 278 999 (15.6) | 48 (13.8) | 298 (15.4) | 326 (18.5) | 1 768 (14.7) |
| 10-12 years | 951 559 (52.9) | 943 516 (52.9) | 165 (47.3) | 1 005 (51.8) | 889 (50.4) | 6 094 (50.7) |
| >12 years | 567 369 (31.5) | 561 887 (31.5) | 136 (39.0) | 636 (32.8) | 549 (31.1) | 4 161 (34.6) |
| **Fathers** |  |  |  |  |  |  |
| Age at time of birth (years ± SD): | 32.2 (6.0) | 32.2 (6.0) | 33.0 (6.4) | 33.1 (6.4) | 32.7 (6.4) | 32.9 (6.1) |
| Country of birth |  |  |  |  |  |  |
| Sweden | 1 529 779 (85.0) | 1 516 229 (85.0) | 293 (84.0) | 1 635 (84.3) | 1 435 (81.4) | 10 187 (84.7) |
| Nordic (except Sweden)* | 46 125 (2.6) | 45 754 (2.6) | 10 (2.9) | 34 (1.8) | 47 (2.7) | 280 (2.3) |
| Other European | 29 296 (1.6) | 29 062 (1.6) | 3 (0.9) | 39 (2.0) | 24 (1.4) | 168 (1.4) |
| Outside Europe | 195 277 (10.9) | 193 357 (10.8) | 43 (12.3) | 231 (11.9) | 258 (14.6) | 1 388 (11.5) |
| Level of highest education<sup>†</sup> |  |  |  |  |  |  |
| ≤ 9 years | 322 059 (17.9) | 319 360 (17.9) | 59 (16.9) | 333 (17.1) | 347 (19.7) | 1 960 (16.3) |
| 10-12 years | 953 908 (53.0) | 945 395 (53.0) | 169 (48.4) | 1 014 (52.3) | 901 (51.1) | 6 429 (53.5) |
| >12 years | 524 510 (29.1) | 519 647 (29.1) | 121 (34.7) | 592 (30.5) | 516 (29.2) | 3 634 (30.2) |
| **Socioeconomy** |  |  |  |  |  |  |
| Total family income/individual‡ |  |  |  |  |  |  |
| Percentile 0-20 | 309 019 (17.2) | 306 259 (17.2) | 58 (16.6) | 335 (17.3) | 337 (19.1) | 2 030 (16.9) |
| Percentile 20-40 | 345 210 (19.2) | 342 078 (19.2) | 59 (16.9) | 384 (19.8) | 333 (18.9) | 2 356 (19.6) |
| Percentile 40-60 | 367 768 (20.4) | 364 453 (20.4) | 71 (20.3) | 408 (21.0) | 358 (20.3) | 2 478 (20.6) |
| Percentile 60-80 | 384 026 (21.3) | 380 571 (21.3) | 87 (24.9) | 374 (19.3) | 345 (19.6) | 2 649 (22.0) |
| Percentile 80-100 | 394 454 (21.9) | 391 041 (21.9) | 74 (21.2) | 438 (22.6) | 391 (22.2) | 2 510 (20.9) |
| **Children** |  |  |  |  |  |  |
| Sex |  |  |  |  |  |  |
| Female | 875 117 (48.6) | 867 310 (48.6) | 138 (39.5) | 829 (42.8) | 924 (52.4) | 5 916 (49.2) |
| Male | 925 360 (51.4) | 917 092 (51.4) | 211 (60.5) | 1 110 (57.3) | 840 (47.6) | 6 107 (50.8) |
| SGA |  |  |  |  |  |  |
| <3rd percentile | 42 181 (2.3) | 41 223 (2.3) | 12 (3.4) | 143 (7.4) | 147 (8.3) | 656 (5.5) |
| Missing | 6378 (0.4) | 6221 (0.3) | 7 (2.0) | 43 (2.0) | 20 (1.0) | 87 (0.7) |
| Gestational age |  |  |  |  |  |  |
| >36 w | 1 711 325 (95.0) | 1 697 498 (95.1) | 318 (91.1) | 1 721 (88.8) | 1 451 (82.2) | 10 337 (86.0) |
| <37 w | 87 045 (4.8) | 84 824 (4.8) | 31 (8.9) | 209 (10.8) | 310 (17.6) | 1 671 (13.9) |
| Missing | 2 107 (0.1) | 2 080 (0.1) | 0 (0) | 9 (0.5) | 3 (0.2) | 15 (0.1) |
| Congenital anomalies |  |  |  |  |  |  |
| No | 1 614 313 (89.7) | 1 603 019 (89.8) | 170 (48.7) | 1 023 (52.8) | 1 118 (63.4) | 8 983 (74.7) |
| Major non-cardiac | 102 327 (5.7) | 98 459 (5.5) | 153 (43.8) | 828 (42.7) | 546 (31.0) | 2 341 (19.5) |
| Non-cardiac | 186 164 (10.3) | 181 383 (10.2) | 179 (51.3) | 916 (47.2) | 646 (36.6) | 3 040 (25.3) |
| **Follow up (MCA excl)** |  |  |  |  |  |  |
| Not in school registry (%) |  | 2.2 | 9.2 | 6.0 | 6.3 | 3.8 |
| Approved math. (%) |  | 90.6 | 81.1 | 83.2 | 84.6 | 87.8 |
| Approved Swedish (%) |  | 93.4 | 85.2 | 87.4 | 88.0 | 91.2 |
| Approved English (%) |  | 92.7 | 84.2 | 86.9 | 86.4 | 90.6 |

Values are numbers (percentages) unless stated otherwise.
\*Finland, Norway, Denmark, and Iceland.
† Highest educational level attained at time of childbirth in number of years of completed education of both parents into; up to 9 years (primary school), 10 to 12 years (secondary school), and more than 12 years (higher education or university). See supplementary Figure 2 for an overview of the Swedish school system.
‡ Individualized disposable family income reflects the financial situation of a household after accounting for all sources of income (including social benefits) for a family, deduction of all outgoing expenses (including taxes), and adjustment for family size. It is updated yearly by Statistics Sweden and is based on mandatory reporting. We extracted the individualized family income at the time of the child's birth and, to avoid misclassification bias because of inflation, coded this into quintiles in relation to year of ascertainment.
Abbreviations: CHD, Congenital Heart Disease; UVH, Univentricular heart disease; SGA<sup>42</sup>, Small for Gestational Age; w, week; MCA, Major non-cardiac congenital anomaly; math., mathematics.

### Congenital heart disease and scholastic performance in adolescence

Overall, children with CHD had worse scholastic performance than did the total population, a difference that was stable after adjusting for sex, birth year, maternal age, paternal age, maternal birth country, maternal education, paternal education, and disposable income/family member. The primary outcome measure, eligibility to USS, and children without non-cardiac MCA, under the assumption that those without any grade were deemed ineligible, demonstrated a linear association with the complexity of CHD, **Figure 1**. In the entire group, non-cardiac CA included, the greatest disparity was shown for children with a severe CHD, RR 3.37 (95% confidence interval 3.11-3.50) of not being eligible, probably explained by CHD-associated MCAs such as Down syndrome and 22q11 in this group, **Figure S3**.

**Figure 1.**
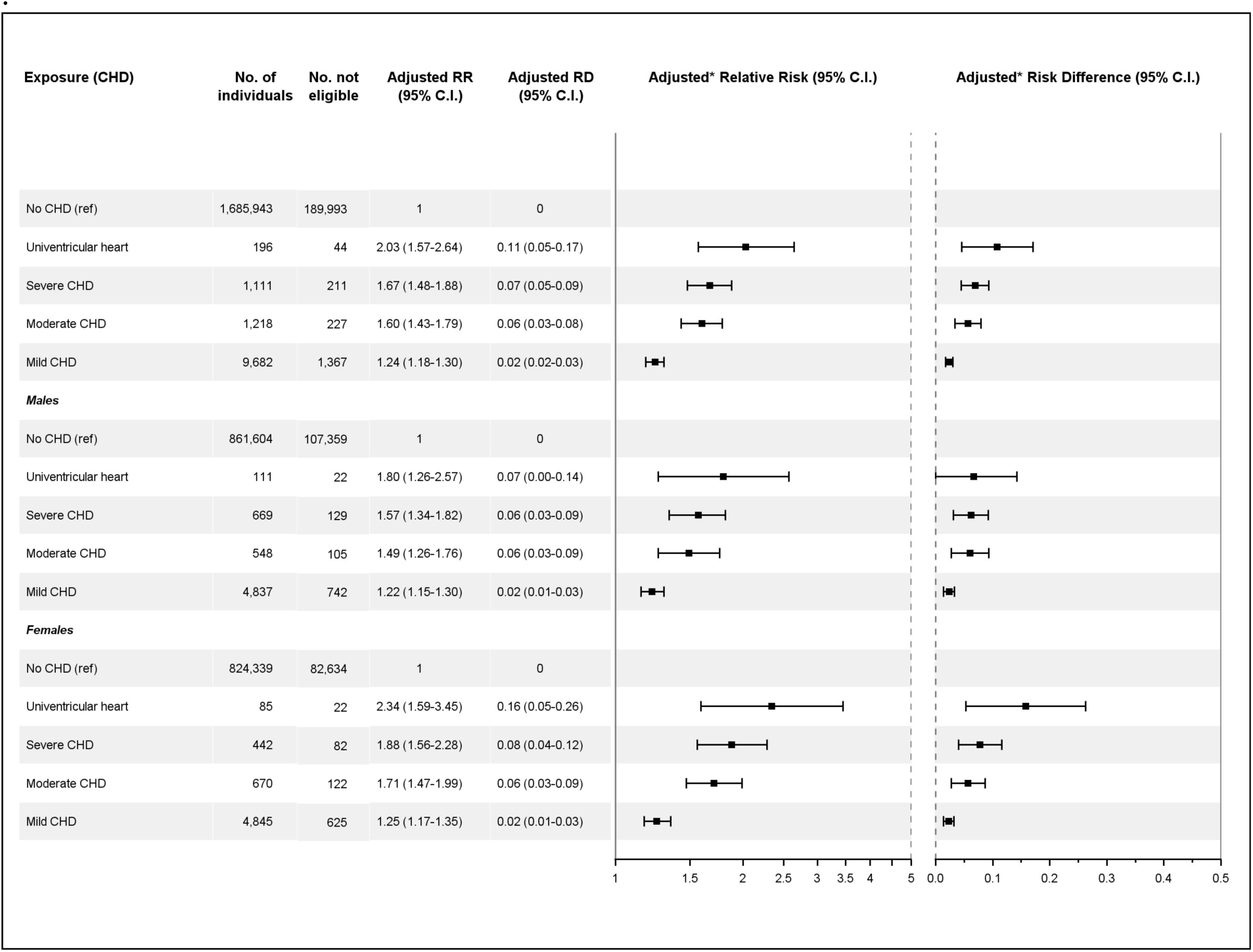
The risk of not being eligible for USS at 16 years of age in relation to the complexity of CHD, MCA excluded. Data were analyzed using multivariate Poisson regression models, adjusted for sex, birth year, maternal age, paternal age, maternal birth country, paternal birth country, maternal education, paternal education, and disposable income/family member. Data are presented as RR with 95% confidence intervals and RD as a percentage (%). Missing grades were imputed as not passing. Abbreviations: USS, Upper secondary school; MCA, Major non-cardiac congenital anomaly; CHD, Congenital Heart Disease; No, Number; RR, Relative Risk; RD, Risk Differences; C.I., Confidence Interval; Ref, Reference group.

Boys and girls demonstrated comparable eligibility for USS, **Table S3**.

Eligibility to USS showed a similar significant trend with non-imputed grades in adjusted and non-adjusted models, **Figure S4** and **Table S3**.

As minor CA did not affect the results, they were retained in the following analysis to improve power, **Table S3**.

The complexity of CHD was associated with the risk of failing core subjects in 9^th^ grade, with an equally elevated risk across all three core subjects, **Table 1**. The probability of achieving a higher grade than pass in the core subject was also hampered in all children with CHD after adjustments, except for the moderate cases, and was borderline significant for UVH cases in mathematics, **Table S4.**

Elucidating the broader impact of CHD on the total grade point in 9^th^ grade, we observed a consistent trend similar to that seen in core subjects. When excluding MCA and stratifying on gender, total grade sum were affected by both the complexity of CHD and gender, where males with UVH had an average just above passed grades in all subjects, but girls with UVH had somewhat higher total grade sum, as shown in **Table 2**.

**Table 2.** Total grade sum for children with or without CHD, MCA excluded.

| **Gender** | **CHD Status** | **Total Crude** | **Total Adjusted\*** | **P value for trend** |
| --- | --- | --- | --- | --- |
| **All** |  |  |  |  |
|  | No CHD | 211 (211-211) | 211 (211-211) |  |
|  | UVH | 178 (168-187) | 174 (165-183) |  |
|  | Severe CHD | 193 (189-198) | 193 (189-197) | p<0.001 |
|  | Moderate CHD | 197 (193-201) | 196 (192-199) |  |
|  | Mild CHD | 208 (207-210) | 204 (202-205) |  |
| **Male** |  |  |  |  |
|  | No CHD | 199 (199-199) | 199 (199-199) |  |
|  | UVH | 178 (165-191) | 168 (157-180) |  |
|  | Severe CHD | 189 (184-194) | 186 (182-191) | p<0.001 |
|  | Moderate CHD | 186 (180-192) | 186 (180-191) |  |
|  | Mild CHD | 196 (194-198) | 192 (190-194) |  |
| **Female** |  |  |  |  |
|  | No CHD | 223 (223-223) | 223 (223-223) |  |
|  | UVH | 177 (162-192) | 178 (164-191) |  |
|  | Severe CHD | 200 (194-207) | 197 (191-203) | p<0.001 |
|  | Moderate CHD | 205 (200-211) | 206 (202-211) |  |
|  | Mild CHD | 221 (219-223) | 216 (214-217) |  |

Data are presented as the total grade sum with a 95% confidence interval. Imputation of no grade in a subject as 0 grade points. All individuals and stratification by sex are presented.
\*Adjusted for sex, birth year, maternal age, paternal age, maternal birth country, maternal education, paternal education, and disposable income/family member.
Abbreviations: Total, total grade sum; CHD, Congenital Heart Disease; CI, Confidence Interval; UVH, Univentricular heart disease; MCA, major non-cardiac congenital anomaly.

### School performance by SGA and GA

SGA and prematurity, individually, nearly doubled the risk of being ineligible for USS in all children with CHD, with RR 2.34 (95% confidence interval 2.04-2.69) and 1.81 (95% confidence interval 1.64-1.99), respectively. However, when subgrouping children in complexity of CHD, significant differences remained for moderate and mild cases without MCAs, compared to children with the same complexities who were not premature or SGA. A non-significant additional risk of being SGA or preterm was observed for UVH and severe CHD; however, these cases were few, which may have led to underpowering in these groups, **Figure 2**.

**Figure 2.**
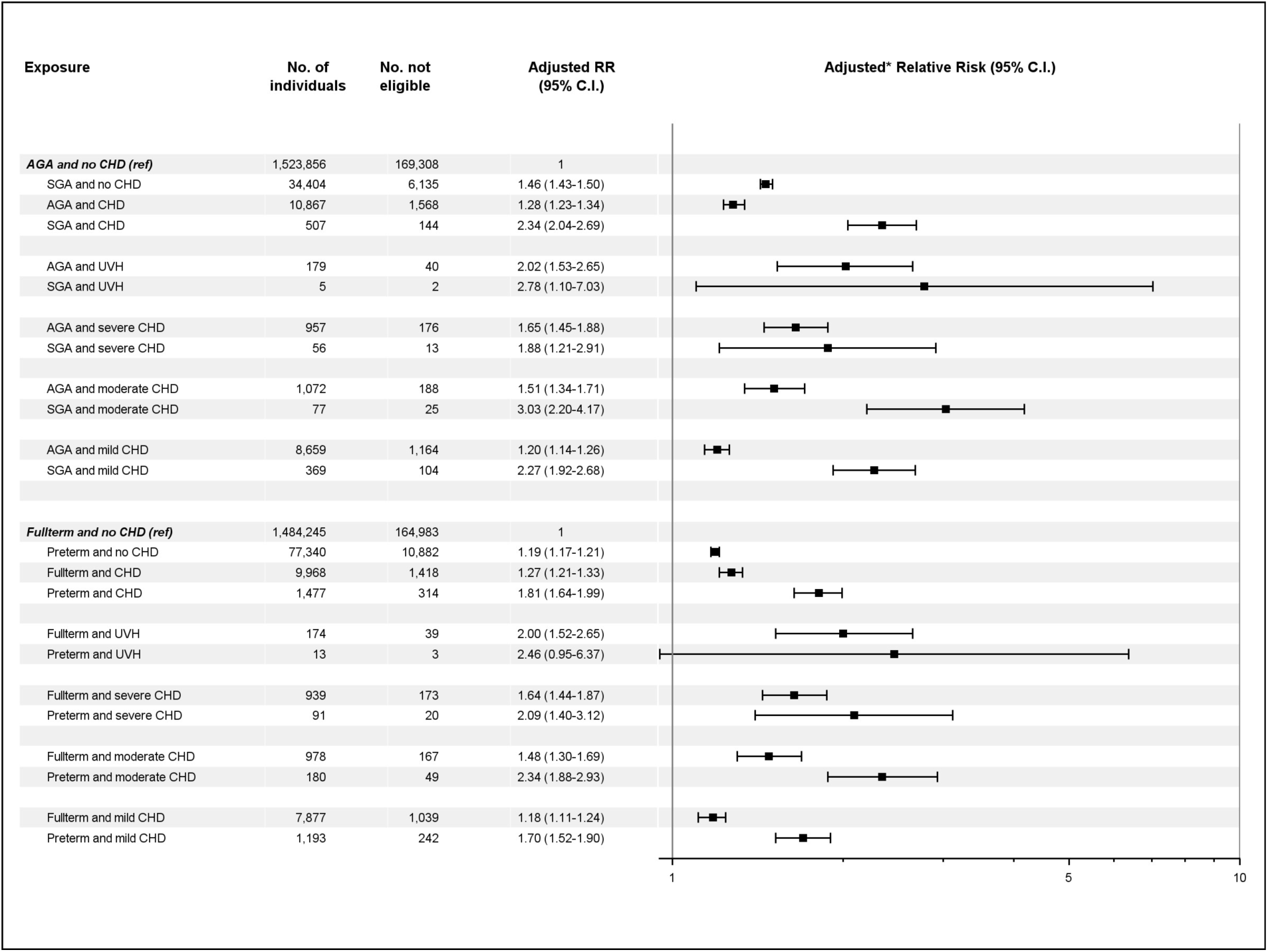
Additive relative risk of being SGA (<3SD) or preterm (<37GA) in relation to the complexity of CHD and ineligibility for USS. Data were analyzed using multivariate Poisson regression models, adjusted for sex, birth year, maternal age, paternal age, maternal birth country, paternal birth country, maternal education, paternal education, and disposable income/family member. Children with a MCA were excluded. Data are presented as RR with 95% confidence intervals and RD as a percentage (%). Missing grades were imputed as not passing. Abbreviations: USS, Upper secondary school; SGA, Small for Gestational Age^42^; GA, Gestational Age; CHD, Congenital Heart Disease; RR, Relative Risk; MCA, Major non-cardiac congenital anomaly; C.I., Confidence Interval; AGA, appropriate for gestational age; UVH, Univentricular heart disease; Ref, reference.

### Eligibility for higher education by period

In our exploratory analysis, stratifying by surgical eras based on birth year, before (1987-1991), during (1992-1996), and after the centralisation of pediatric heart surgery (1997-2005), did not reveal any change in the prevalence of eligibility for higher education over time across all complexities of CHD, **Figure 3**.

**Figure 3.**
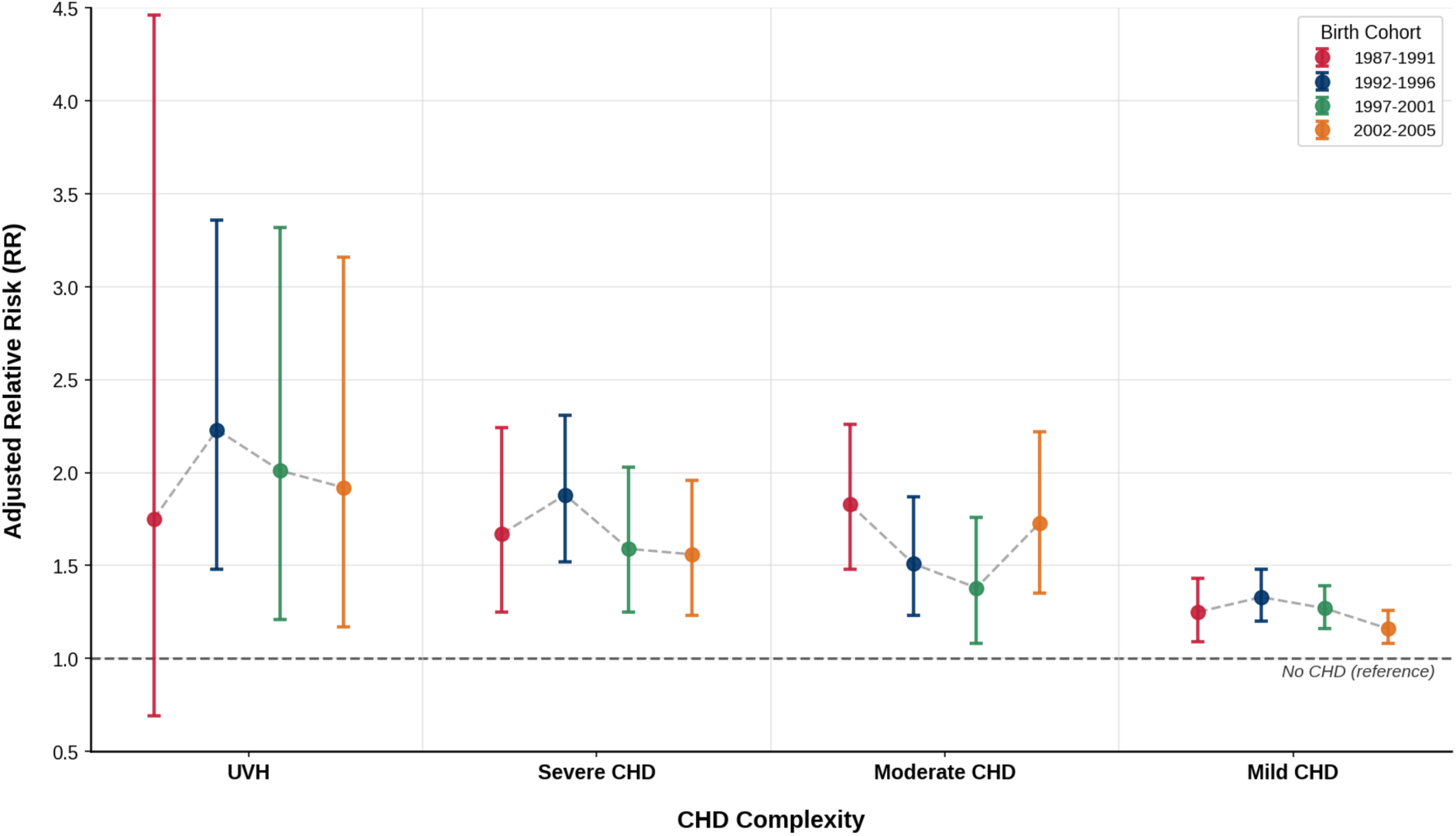
Relative risk of not being eligible for USS by birth cohorts in children with CHD, MCA excluded. The birth cohort from 1987–1991 represents the period before, 1992–1996 during, and 1997–2001 and 2002–2005 after the centralisation of pediatric heart surgery in Sweden. Data are presented as RR with a 95% confidence interval. Missing grades were imputed as not passing. No CHD serves as the reference. Adjusted for sex, birth year, maternal age, paternal age, maternal birth country, paternal birth country, maternal education, paternal education, and disposable income/family member. Abbreviations: USS, Upper secondary school; CHD, Congenital Heart Disease; UVH, Univentricular heart disease; MCA, major non-cardiac congenital anomaly; RR, Relative Risk.

### School performance in relation to siblings

Sibling comparisons revealed the same risk of not being eligible for USS across the complexities of CHD, as when compared to the general population, after excluding MCA. For the entire group, including all CAs, differences between siblings were somewhat more pronounced than to the general population. This is likely due to the inherent neurodevelopmental impairments associated with many of the genetic aberrancies also causing CHD^26^, **Table S5**. Stratification by gender revealed a comparable risk of ineligibility across all levels of complexity of CHD. However, the analysis was limited by wide CIs, attributed to attrition, thereby reducing statistical power.

## Discussion

In this comprehensive nationwide cohort of 1,8 million children, all CHD was associated with poorer scholastic performance in adolescence. Notably, the extent of academic impairment increased with the complexity of the CHD. Outcomes worsened with MCAs, prematurity, and SGA. The findings remained consistent across sibling comparisons by sex and birth cohorts.

Impaired cognitive outcomes are the most common self-reported disabilities in adults with CHD^7^, affecting their socioeconomic status^44^ and increasing mortality^45^. Unlike previous studies, which are numerous and often conducted in childhood, showing impaired intelligence or executive functions through single tests^21,46,47^, our study demonstrates the combined effect of cognitive abilities and personality on grades in adolescence, which presumably better predicts life outcomes^48^. Prior research on academic performance has yielded inconsistent results^19-24^. Studies in early childhood often show less difference in academic achievement in children with CHD compared to healthy peers^22^, potentially overestimating their later performance^49,50^. Long-term data from large groups indicate lower educational attainment, with one study showing a declining trend over the past two decades^21^. However, these studies either lacked socioeconomic adjustments or failed to adequately account for the need for surgery^19,21,22^.

Our findings reveal that poorer academic outcomes were consistent over time across severity groups, indicating that increased survival hasn’t led to improved neurodevelopment. The current study also broadened the insight into academic outcomes, looking not only at core subjects, but at the composite grade of all subjects, which has previously been unrecognized. The trend of lower achievement in core and all subjects is consistent with increasing CHD complexity. Although socioeconomic factors are recognized as major determinants of neurocognitive outcomes in children with CHD^45^, our results remained robust even after adjusting for socioeconomic status. This resilience may partly reflect the mitigating effect of Sweden’s universal, publicly funded healthcare and education systems.

We also found that children with moderate and mild non-surgical CHD had slightly impaired academic performance, in contrast with some previous studies on neurodevelopmental outcomes^51,52^. Growing knowledge of genetic anomalies linked to CHD and neurological disabilities, together with the known hemodynamic effects of CHD during fetal life, may explain these findings^10,26^. Our results demonstrating that prematurity adds additional negative effects on educational outcomes align with recent research^21^, suggesting a potential ‘double-hit’ effect on neurodevelopment as previously outlined^53,54^. In the general population, late preterm birth is associated with worse neurodevelopmental outcomes^55^, though this is not mirrored in their academic performance^37^.

This study justifies the AHA’s risk stratification framework, as neurodevelopmental outcomes associated with CHD complexity and prematurity are reflected in academic performance during adolescence. However, we also find that SGA affects academic achievement, as previously shown to worsen neurodevelopment^17^, but is not identified as a risk factor in the AHA statement^4^.

Recognition of the reduced likelihood of higher educational attainment among children with CHD, particularly those with more complex conditions, underscores the need for strategies to mitigate these disparities. Recently, a neurodevelopmental program in infancy has shown short-term cognitive gains^56^. In contrast, an intervention aimed at improving cognition in childhood has not yielded similar benefits in children with CHD^57^. Moreover, screening for neuropsychiatric comorbidities in children with CHD could be challenging, as they may not fulfill criteria for any single neuropsychiatric diagnosis, yet still experience a range of difficulties across the neuropsychiatric spectrum^4^. Acknowledging these challenges is essential to ensure that children with CHD receive appropriate support from child psychiatric, educational, and social services^58^. Although the overall burden of neuropsychiatric disorders in the CHD population remains to be fully elucidated, emerging evidence indicates a significantly increased prevalence^5^. The current data justifies the change in obstetrical practice with increasing gestational age at delivery^39^. For clinicians, these findings provide additional insight to support antenatal counselling. Genetic aberrancies associated with CHD significantly influence school achievements, and genetic testing should be empowered.

### Strengths and limitations of this study

A major strength of this study is its large, population-based design set in a contemporary context. Additionally, the Swedish population is diverse, enhancing generalizability^59^. The Swedish registers provide extensive, high-quality data, allowing us to examine nearly all Swedish children with CHD and their academic achievements, while adjusting for socioeconomic factors. Our imputation of grades uncovered greater disparities between children with CHD and the overall population. Significant differences remained in both unadjusted models and those incorporating non-imputed grades, thereby establishing credibility. Our inclusion criteria, based on diagnosis, surgical procedure codes, and timing of surgery, may have misclassified some children with CHD as controls, potentially underestimating associations^60^. Our proportion of children with CHD is well in line with reported prevalences^1,2^, although recognizing an increasing trend amongst mild CHD cases, such as patent ductus of prematurity, which was not considered a CHD in the current study. Moreover, 2.0% of the population were excluded from the study cohort due to missing socioeconomic data, and 5.1% were excluded due to multiparty, emigration, or death. However, our reported number of UVHs aligns with a recent publication on long-term survival after univentricular palliation in Sweden^35^.

The exact number of children requiring special education in the current population remains unknown, and changes to the grading system in 2012, despite adjustments, introduce limitations. Another limitation is that the number of early-term deliveries declined in Sweden between 2006 and 2014^39^, which could result in improved school performance in the current cohort, since even early-term delivery has been linked to adverse neurodevelopmental outcomes, especially in children with UVH^53,54^.

### Conclusion

This extensive population-based study found that there is an inherent risk of adverse academic outcomes in adolescence in all children with CHD that increases with complexity. Fortunately, we found that most children with CHD will be eligible for higher education. Still, healthcare personnel and policymakers need to be aware of the children’s challenges. Further studies should focus on efficient interventions to ameliorate cognitive outcomes in children with CHD, particularly since they comprise a significant and growing population.

## Data Availability

Data not available due to GDPR.

## Non-standard Abbreviations and Acronyms

AHA: American Heart Association
CHD: congenital heart disease
GA: gestational age
ICD: International Classification of Diseases
K6, KVÅ/KKÅ: Classification of Health Care Interventions and Surgical Procedures
MBR: Medical Birth Register
MCA: major non-cardiac congenital anomalies
NPR: National Patient Registry
SGA: small for gestational age
USS: upper secondary school
UVH: univentricular heart

## Acknowledgments

*Patient and public involvement*

The parental organization for children with CHD in Sweden advised on which outcomes are important to patients. Still, they were not involved in the conduct, analysis, or reporting of the research.

## Funding

SES received funding from the Swedish Heartchild Foundation, VS from the Donation Fund for Research on Neurodevelopmental Disorders at Karolinska University Hospital, GB from the Swedish Heart Lung Foundation and UÅ from the Regional ALF Grants Region Östergötland. The funders had no role in any aspect of the study, and all authors acted independently from the study sponsors.

## Disclosures

None

## Supplemental Material

Table S1-S5

Figure S1-S4

## Notes

### Competing Interest Statement

The authors have declared no competing interest.

### Clinical Trial

This was not a clinical trial.

### Author Declarations

The study was conducted in accordance with the Helsinki Declaration and approved by the Regional Ethical Review Board in Stockholm, Sweden, DNR: 2020-05516, 2021-05958-02, 2022-05648-02 and 2024-00060-01.

